# Structural and diffusion weighted brain imaging biomarkers for ADHD and its symptomology in very young (4–7-year-old) children

**DOI:** 10.1101/2021.09.23.21263990

**Authors:** Ilke Öztekin, Dea Garic, Mark A. Finlayson, Paulo A. Graziano, Anthony Steven Dick

## Abstract

The current study aimed to identify the key neurobiology of Attention-Deficit/Hyperactivity Disorder (ADHD), as it relates to ADHD diagnostic category and symptoms of hyperactive/impulsive behavior and inattention. To do so, we adapted a predictive modeling approach to identify the key structural and diffusion weighted brain imaging measures, and their relative standing with respect to teacher ratings of executive function – EF (measured by the Metacognition Index of the Behavior Rating Inventory of Executive Function– BRIEF), negativity and emotion regulation – ER (measured by the Emotion Regulation Checklist, ERC), in a critical young age range (ages 4 to 7, mean age 5.52 years, 82.2% Hispanic/Latino), where initial contact with educators and clinicians typically take place. Teacher ratings of EF and ER were predictive of both ADHD diagnostic category and symptoms of hyperactive/impulsive behavior and inattention. Among the neural measures evaluated, the current study identified the critical importance of the largely understudied diffusion weighted imaging measures for the underlying neurobiology of ADHD and its associated symptomology. Specifically, our analyses implicated the inferior frontal gyrus, the pericallosal sulcus, and the caudate as critical predictors of ADHD diagnostic category and its associated symptomology, above and beyond teacher ratings of EF and ER. Collectively, the current set of findings have implications for theories of ADHD, the relative utility of neurobiological measures with respect to teacher ratings of EF and ER, and the developmental trajectory of its underlying neurobiology.

## INTRODUCTION

Attention-Deficit/Hyperactivity Disorder (ADHD) is a developmental disorder that affects over 7% of children worldwide (Wolraich et al., 2019). The etiology of ADHD at the neurobiological level is not well-established, although there is a general consensus that frontal, parietal, basal ganglia, and cerebellar regions of the dopaminergic system, as well as some of the fiber pathways facilitating functional interactions among these brain regions, are affected. These regions also support cognitive and affective processes that are often impaired in children with ADHD, such as executive function (EF) and emotion regulation (ER). EF, which is an umbrella term for the cognitive processes such as attentional control, inhibitory control, and performance monitoring necessary for voluntary control of behavior, is a prominent, though not universal, feature of ADHD pathophysiology (Barkley, 1997; Sergeant, 2000; Sonuga-Barke, 2002). With respect to the neurobiological markers that support EF, prior research has pointed to functional interactions among lateral frontal, inferior frontal/insular (Aron et al., 2004), medial frontal/anterior cingulate/pre-SMA (Aron et al., 2004; Bunge & Wright, 2007; Fedota et al., 2014; Miller & Cohen, 2001; Rushworth et al., 2005), lateral parietal (Corbetta & Shulman, 2002), and dorsal striatal (Morein-Zamir & Robbins, 2015) regions of the brain (Hart et al., 2014).

ER, or dysregulation, is also a prominent behavioral profile in pediatric ADHD (Barkley & Fischer, 2010; Graziano & Garcia, 2016; Karalunas et al., 2019; Shaw et al., 2003). Notably, a meta-analysis carried out by Graziano and Garcia (Graziano & Garcia, 2016) demonstrated that ER deficits were independent of co-occurring conduct problems and were in similar or greater magnitude with respect to the EF deficits observed among children with ADHD. The importance of such ER deficits are in line with theories of ADHD that have suggested EF-related effects might not emerge in the young age range evaluated in the current study, and that subcortical regions that support ER might be crucially important for the early onset of ADHD (Halperin & Schulz, 2006). Indeed, findings from recent neural investigations (Hoogman et al., 2020; Oztekin et al., 2021) provide support for the contention that a primary focus on the neurobiology of EF alone might not be sufficient, and not sensitive to the heterogeneity of developmental trajectories of ADHD, especially in the critical young age range assessed in the current study. Thus, when considering the predictive utility of neurobiological markers for ADHD, it appears important and necessary to conjointly consider the underlying neural mechanisms that support EF and ER. The neurobiology of ER processes constitutes functional interactions of the medial and dorsal prefrontal cortical structures, anterior cingulate cortex, and medial orbitofrontal cortex with amygdala (Albaugh et al., 2013; Beauregard et al., 2001; Davidson & Slagter, 2000; Lane & McRae, 2004). The major fiber pathways supporting connectivity among these regions are the *uncinate fasciculus* (connecting amygdala with lateral and orbital and medial prefrontal cortex, e.g., (Pacheco et al., 2009) and the *cingulum* (connecting amygdala with medial frontal and cingulate, e.g., see (Jones et al., 2013).

In line with this contention, the current study adapted a predictive modeling approach that leverages machine learning to evaluate the utility of target measures of EF and ER (neurobiology of EF and ER, along with teacher ratings of EF and ER) in predicting the early onset of ADHD in children ages 4 to 7. Specifically, the present study evaluates the incremental predictive value of the target neurobiology in comparison to the target measures of teacher ratings of EF and ER in this young age range as it relates to 1) diagnostic classification of ADHD, and 2) ADHD symptomology of inattention and hyperactive/impulsive behavior dimensionally (see Petrovic & Castellanos, 2016; Woo et al., 2017 for a critical overview of the importance of dimensional approaches in clinical neuroscience).

Our assessment of neurobiology using a machine learning approach leverages structural brain imaging measures, as well as diffusion-weighted imaging measures of neurite density and fractional anisotropy in three fiber pathways of importance to ADHD in this ange range (Cooper et al., 2015; Frodl & Skokauskas, 2012; Garic et al., 2019; Graziano et al., 2021; Jones et al., 2013; Konrad & Eickhoff, 2010; Nagel et al., 2011; Peterson et al., 2011; van Ewijk et al., 2012), namely the frontal aslant tract (FAT), cingulum, and the uncinate fasciculus (UF). In addition to assessing the predictive performance of the target measures, we further evaluated their relative importance. Our main hypothesis was that the identified critical neurobiology for predicting ADHD diagnostic category would extend to regions important for both EF and ER, and that the critical neurobiology for predicting diagnostic category would also hold a reliable relationship with dimensional symptoms of hyperactive/impulsive behavior and inattention. Our third goal was to further identify the neurobiology that contributes to differentiating ADHD diagnostic category and symptomology above and beyond the utility provided by teacher ratings alone.

Our study leverages a unique sample, the ADHD Heterogeneity of Executive Function and Emotion Regulation Across Development (AHEAD) study, which aims to characterize the heterogeneity of well-established predictors of ADHD among young children (ages 4-7, mean age 5.52, 82.2% Hispanic/Latino) across multiple levels of analysis. Notably, AHEAD is among the first to scan children with ADHD as young as 4-7 years, where early diagnosis is critically important (see also Jacobson et al., 2018; Rosch et al., 2018; for notable prior studies with an older age range focus, see Fair et al., 2012; Karalunas et al., 2014; Karalunas et al., 2019; Qureshi et al., 2016; Qureshi et al., 2017). With its multimodal imaging approach, it further provides a unique opportunity to evaluate structural brain imaging and diffusion-weighted imaging metrics conjointly, and how they compare to critical measures of teacher ratings (for both EF and ER) of high importance in this very young age range.

## MATERIALS AND METHODS

### Participants and Recruitment

Children and their caregivers were recruited from local schools and mental health agencies via brochures, radio and newspaper ads, and open houses/parent workshops. Legal guardians contacted the clinic and were directed to the study staff for screening questions to determine eligibility. For the ADHD sample, if the parent (1) endorsed clinically significant levels of ADHD symptoms (six or more symptoms of either Inattention or Hyperactivity/Impulsivity according to the DSM-5 OR a previous diagnosis of ADHD), (2) indicated that the child is currently displaying clinically significant academic, behavioral, or social impairments as measured by a score of 3 or higher on a seven-point impairment rating scale (Fabiano et al., 2006), and (3) were not taking any psychotropic medication, the parent and child were invited to participate in an assessment to determine study eligibility. For the typically developing sample, if the parent (1) endorsed less than 4 ADHD symptoms (across either Inattention or Hyperactivity/Impulsivity according to the DSM-5), (2) less than 4 Oppositional Defiant Disorder (ODD) symptoms, and (3) indicated no clinically significant impairment (score below 3 on the impairment rating scale), the parent and child were invited to participate in an assessment to determine study eligibility. Participants were also required to be enrolled in school during the previous year, have an estimated IQ of 70 or higher, have no confirmed history of an Autism Spectrum Disorder, and be able to attend an 8-week summer treatment program prior to the start of the next school year (ADHD groups only). Due to the young age of the sample, only disruptive behavior disorders were extensively examined for diagnostic purposes.

During intake, ADHD diagnosis (and comorbid disruptive behavior disorders) was assessed through a combination of parent structured interview (Shaffer et al., 2000) and parent and teacher ratings of symptoms and impairment (Fabiano et al., 2006) as is recommended practice (Pelham et al., 2005). Specifically, the DBD rating scales and diagnostic interview were combined using an “or rule,” which identifies the presence of a symptom if endorsed by either informant while clinically significant problems at home and school were defined by at least a “3” on a “0 to 6” impairment rating scale (Bird et al., 1992; Sibley et al., 2016). Dual Ph.D. level clinician review was used to determine diagnosis. Of relevance to the current study, the BRIEF rating scale was not used for any diagnostic purposes. The analyses reported in this paper included 180 children who had data available from all the behavioral and neural target measures of interest, described further below. Our sample comprises of 85 children with ADHD (47.2%), and 95 (52.8%) typically developing children. Among the participants with ADHD, 59 (69.4%) had comorbid ODD diagnosis. Parental consent and assent were obtained in accordance with the Office of Research Integrity at Florida International University.

### Target Measures

#### Teacher Ratings of Executive Function and Emotion Regulation

We used the Emergent Metacognition Index (MCI) *t*-score from the Behavior Rating Inventory of Executive Function, both Child and Preschool versions (BRIEF Child & BRIEF-P; Gioia, Espy & Isquith, 2003, Cronbach’s alpha .99) for our measure of teacher ratings of executive function. The MCI is thought to reflect the ability to maintain information and/or activities in working memory, as well as to plan and organize problem-solving approaches. In the BRIEF Preschool, the MCI is composed of the Working Memory and Plan/Organize scales. In the BRIEF Child, the MCI is composed of the Initiate, Working Memory, Plan/Organize, Organization of Materials, and Monitor scales. We used the Emotion Regulation and Negativity z-scores from the Emotion Regulation Checklist (ERC; Shields & Cicchetti, 1997, Cronbach’s alpha .99) as our target measures for teacher ratings of emotion regulation in our sample.

#### Neural Measures

Our neurobiological measures of interest include structural brain imaging measures of cortical thickness, volume, surface area, and curvature, as well as diffusion-weighted imaging measures of neurite density and fractional anisotropy in the three fiber pathways assessed, namely the frontal aslant tract, the cingulum, and the uncinate fasciculus bilaterally. Due to our theoretical focus on EF and ER, our paper primarily focuses on the neurobiology of EF and ER (cortical, subcortical volume and neurite density, see Figure 1). However, we also report machine learning models on the whole brain measures in Supplementary Materials (interested readers can refer to Supplementary Table 1 for performance metrics derived from whole brain measures).

**Figure 1.**
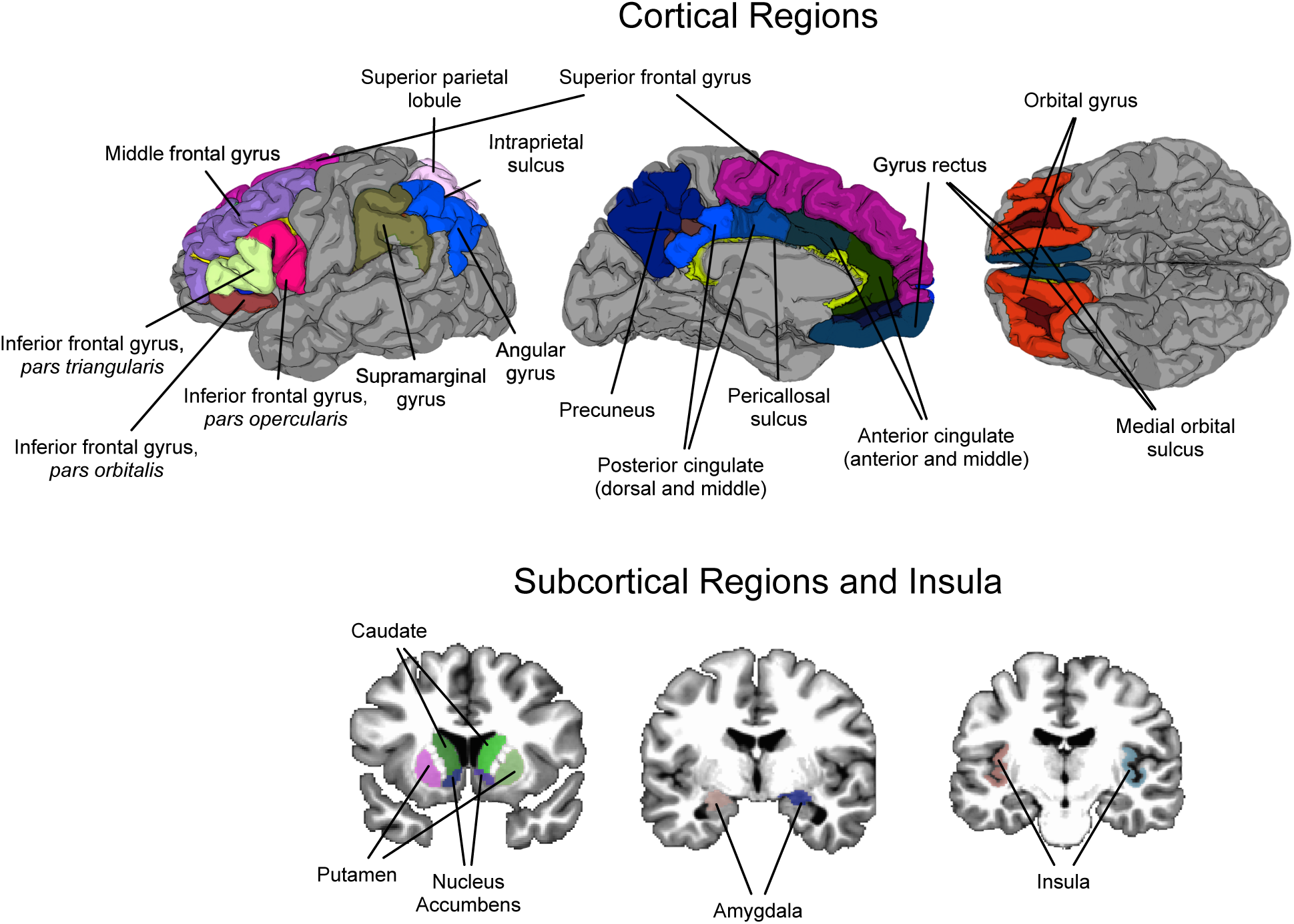
Illustration of the target neurobiology assessed in our study.

All imaging was performed using a research-dedicated 3 Tesla Siemens MAGNETOM Prisma MRI scanner (V11C) with a 32-channel coil located on the university campus. Children first completed a preparatory phase using realistic mock scanner in the room across the hall from the magnet. Here they were trained to stay still, and were also acclimated to the enclosed space of the magnet, the back projection visual presentation system, and to the scanner noises (in this case, presented with headphones). When they were properly trained and acclimated, they were moved to the magnet. In the magnet, during the structural scans, children watched a child-friendly movie of their choice. Ear protection was used, and sound was presented through MRI compatible headphones.

Structural MRI scans were collected using a 3D T1-weighted sequence (axial; 1 x 1 x 1 mm, 7 min 14 sec) with prospective motion correction (Siemens vNAV; Tisdall et al., 2012), according to the Adolescent Brain and Cognitive Development (ABCD) protocol (Hagler Jr et al., 2019). To provide a semi-automated parcellation of the cerebral cortices and volume of subcortical structures, we constructed two-dimensional surface renderings of each participant’s brain using FreeSurfer v6.0 (Dale et al., 1999; Fischl & Dale, 2000). We computed cortical thickness as part of the standard FreeSurfer reconstruction pipeline (Rohde et al., 2004), as this has been shown to have high correspondence to histological measurements of cortical thickness (Yeh et al., 2010).

Diffusion Weighted (DW) scans were acquired via high-angular resolution diffusion imaging (HARDI) with B_0_ EPI distortion correction (EPIC; TR/TE = 4100/88 ms; 1.7 mm x 1.7mm x 1.7mm; 81 slices no gap; 102 diffusion directions: *b* = 500 (6-dirs); 1500 (15-dirs); 2000 (15-dirs); 3000 (76-dirs) s/mm^2^; A-to-P direction (7 m 31 s). Using the TORTOISE DIFFPREP and DRBUDDI software, and DTI_Prep (Rohde et al., 2004), DW images were processed for motion correction, to remove eddy current artifacts, to correct for EPI B0 susceptibility deformations, to conduct B-matrix reorientation, and to co-register the diffusion series with structural MRI.

Initial quality control was accomplished in DTIPrep to complete the following steps: 1) image/diffusion information check; 2) padding/cropping of data; 3) Rician noise removal; 4) slice-wise, interlace-wise, and gradient-wise intensity and motion checking. In this step, inter-slice brightness artifact detection is accomplished via normalized correlation analysis between successive slices. In addition, interlaced correlation analysis is used for detection and removal of “venetian blind” artifacts. An automated kick-out procedure was used to reject and remove bad diffusion acquisitions that met criteria for removal, using the default settings. The number of remaining diffusion scans was used a proxy for movement/scan quality. Eddy current correction was not applied at this phase. Instead, in the second phase, the corrected data were ported from DTIPrep into TORTOISE DIFFPREP, which was used to accomplish motion and eddy current correction. The data were not resampled to a template space, but were kept in the original subject space. In the third phase, FSL topup was used to correct for EPI distortions using the field map, which was collected in the opposite phase encoding direction of the main scan (Andersson et al., 2003; Smith et al., 2004). In the fourth phase, we implemented calculation of the diffusion tensor model in DSI Studio to estimate the eigenvalues reflecting diffusion parallel and perpendicular to each of the fibers along 3 axes (x, y, z). The resulting eigenvalues were then used to compute indices of fractional anisotropy (FA).

### Fiber Tract Identification

Tractography was conducted either using DSI Studio’s built-in tractography atlas (Yeh et al., 2018; for the cingulum and uncinate fasciculus), or (for the frontal aslant) by use of ROI-to-ROI tracking using a modification of the Freesurfer atlas. The DSI Studio built-in tractography atlas was originally created from 840 healthy adults in the HCP840 dataset, and defines white matter regions of interest (ROIs) in the MNI space. The atlas is then non-linearly warped to the native participant space (Yeh et al., 2018). Because we are analyzing a pediatric dataset, each ROI was visually inspected to ensure that any warping to the atlas template did not introduce inaccuracies. Within this atlas the, following two tracts are defined:

#### Uncinate fasciculus (UF)

UF has rostral terminations projecting to the orbital and lateral frontal cortex, to the frontal pole, and to the anterior cingulate gyrus (mainly BAs 10, 11, 32, and 47). The posterior termination in the temporal lobe includes projections through the amygdala, with terminations in the temporal pole (BA 38), uncus (BA 35), and parahippocampal gyrus (BA 30 and 36) (Dick et al., 2013; Holl et al., 2011; Thiebaut de Schotten et al., 2012; Von Der Heide et al., 2013).

#### Cingulum

The cingulum as a whole is composed of a number of smaller short association fiber systems that course in the white matter under the cingulate gyrus. The pathway supports connections to/from lateral and dorsal prefrontal cortex, medial prefrontal cortex and anterior cingulate, insula, parahippocampal gyrus, subiculum, and amygdala (Jones et al., 2013).

Manual ROI-to-ROI tracking was used for the frontal aslant tract, to better isolate fibers terminating/originating in the pre-Supplementary Motor Area (pre-SMA) and posterior inferior frontal gyrus.

#### Frontal Aslant Tract (FAT)

FAT is a monosynaptic fiber pathway connecting the lateral inferior frontal cortex with the medial superior frontal gyrus, and possibily cingulate gyrus. Most of the tract is comprised of fibers connecting the *pars opercularis* with the functionally-defined pre-supplementary motor area (pre-SMA) (Dick et al., 2019). We used a semi-automated approach to define regions of interest (ROIs) for tractography of the frontal aslant. FreeSurfer v6.0 was used for the initial cortical parcellation and cortical segmentation. Next, the Desikan-Killiany Freesurfer atlas (Desikan et al., 2006) for each participant was modified using the procedure specified by Cammoun et al. (Cammoun et al., 2012), originally defined by Hagmann et al. (Hagmann et al., 2008). In this procedure, the larger ROIs from the Freesurfer parcellation are further divided into smaller units (collectively these atlas modifications are called the Lausanne atlases). We used the modification defining 463 total ROIs, which allowed a finer parcellation of the superior frontal gyrus, allowing for a more accurate subdivision of the pre-SMA and SMA. For the current analysis, the fibers passing to/from the pre-SMA and *pars opercularis* were defined. An angular threshold of 40 degrees was employed, with 1,000,000 seeds set as the termination threshold.

### Neurite orientation dispersion and density imaging (NODDI) Metrics

With a multi-shell DWI HARDI acquisition it is possible to quantify tissue microstructure in terms of neurite orientation and density. The Neurite Orientation Dispersion and Density Imaging (NODDI) model is a three-component model that distinguishes the effects on water diffusion in different cellular environments (intra-neurite, extra-neurite, and cerebrospinal fluid (Jespersen et al., 2010; Jespersen et al., 2012; Zhang et al., 2012). The NODDI model allows estimation of the contributions of neurite morphology from the diffusion signal, and such estimates such as neurite density from the NODDI model have been verified with histology in animals (Sato et al., 2017) and pathological findings in humans (Sone et al., 2020). In the present study we focus on the orientation dispersion index (ODI) and the intraneurite volume fraction (INVF) measure, which in gray matter is an index of dendritic and axonal density. In some models this INVF measure is also referred to as the neurite density index (NDI), which is the terminology we will adapt. We computed the ODI and NDI metrics using the Microstructure Diffusion Toolbox (Harms et al., 2017; Harms & Roebroeck, 2018). With the DWI data set registered to the high-resolution anatomy in original subject space, we computed the average NDI for each region of interest defined by our segmentation algorithms.

#### Quality Control of Magnetic Resonance Imaging Scans

Movement artifacts in T1-weighted MRI scans are common, especially in pediatric populations in this age range, and especially in children with ADHD. Fortunately, FreeSurfer is robust to movement-related artifacts, as, except in extreme cases, the program is able to accurately identify intensity differences between white matter and grey matter inherent in the T1-weighted image. In some cases, however, manual intervention is necessary. In this manual intervention, each individual MRI scan is inspected, and in cases where the program does not adequately identify the appropriate regional boundaries, manual edits are employed. We also visually rated each T1-weighted image on a seven-point scale ranging from “Poor = 1” to “Excellent = 4”, with allowances for half-points (e.g., 3.5). For diffusion-weighted imaging scans, we used the number of directions kept as an indicator for scan quality/movement. These quality measures (T1 scan quality for structural imaging measures of cortical thickness, volume, surface area and curvature and number of directions kept for DWI measures of neurite density and fractional anisotropy in the fiber pathways assessed) were used as covariates in data analyses.

### Outcome Measures

Machine learning models assessed ADHD diagnostic category (ADHD present, ADHD absent). Regression analyses assessed ADHD symptomology on a continuum, evaluating symptoms of hyperactive/impulsive behavior and inattention.

### Predictive Modeling Approach

We employed the *scikit-learn* (version 0.23.1, https://scikit-learn.org/stable/) open-source machine learning library for constructing our models. In order to be able to extract feature importance from classifier coefficients, we adapted a Support Vector Machine (Cortes & Vapnik, 1995) classifier with a linear kernel. For each model, target features were scaled using the StandardScaler function in scikit-learn library, which standardizes features by removing the mean and scaling to unit variance. For model validation, we leveraged the built-in cross-validation function of the scikit-learn library. In this approach, the data is split into training and test sets. This is repeated five times, using different portions of the data as training and test. Specifically, in each iteration, the classifier is tested on a portion of the data set that it did not see during training, following the recommended approach in the field (Varoquaux, 2018; Varoquaux et al., 2017). Performance was then evaluated with the commonly employed accuracy scores, as well as F_1_ scores obtained across the cross-validation indices for each model. F_1_ scores are a classification performance metric that is calculated based on the precision (p) and recall (r), 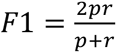. Following the recommended practice in the field, our assessment of statistical significance employed permutation tests (Combrisson & Jerbi, 2015; Noirhomme et al., 2014; Pereira et al., 2009).

Recursive Feature Elimination (RFE). RFE (Guyon et al., 2002) is a recommended feature selection method that has been previously applied in machine learning applications of ADHD (Arbabshirani et al., 2017; Colby et al., 2012; Qureshi et al., 2016; Qureshi et al., 2017; Tan et al., 2017). RFE leverages the coefficients of the classifier in order to select features by recursively considering smaller and smaller set of features. In each iteration, the importance of each feature is derived, and the least important features are pruned from the current set of features. This procedure is recursively repeated on the pruned set until the optimum number of features are selected. Thus, for structural (cortical and subcortical volume) and microstructure (NDI and ODI) measures that contain EF and ER regions, as well as our whole-brain analyses containing all regions of the Destrieux parcellation reported in our Supplementary Materials, we employed recursive feature elimination with cross-validation (RFECV function in scikit-learn library) to identify the optimum set of features that are most informative for predicting ADHD diagnostic category.

### Regression Analyses

To further evaluate the relationship between our target measures and ADHD symptomology, we conducted linear regressions using the OLS function in *statsmodels* library (version 0.11.0) in Python. For each outcome variable (symptoms of hyperactive/impulsive behavior, symptoms of inattention), separate models were run for each set of target measures (teacher ratings of EF and ER, neural measures of volume, neurite density, and fractional anisotropy target pathways— FAT, cingulum, and UF). Age and gender were entered as covariates in all regression analyses. In addition, for analyses that assessed neural measures, whole brain volume and scan quality (T1 quality for structural brain imaging measures, and the number of directions kept for diffusion weighted imaging measures) were included as covariates in the model.

### Overview of Analytical Approach

In summary, our analytical approach adapted the following steps: 1) Use machine learning to determine the relative performance of the target measures in predicting ADHD diagnostic category; 2) For measures that can significantly predict diagnostic category, carry out regression analyses to assess if they can further predict ADHD symptomology (symptoms of hyperactive/impulsive behavior and inattention). RESULTS

### Predicting ADHD diagnostic category

We first assessed the relative predictive utility of the teacher ratings of executive function (BRIEF), emotion regulation (ERC), and the neural measures for predicting ADHD diagnostic category. For this categorical assessment, we trained an SVM to distinguish the TD and ADHD participants. As explained in our Methods section, for neural measures, a feature selection approach that employed recursive feature elimination over the entire set of features was initially adapted to identify the optimum set of target features in predicting ADHD diagnostic category. Below we present our findings across the models. Figure 2 plots the classifier success for predicting ADHD diagnostic category across our primary target models, and Supplementary Table 1 presents the classifier performance metrics for the whole brain measures. Figures 3-5 represent the selected EF and ER regions for our neural metrics, along with their feature importance rankings derived from the classifier coefficients.

**Figure 2.**
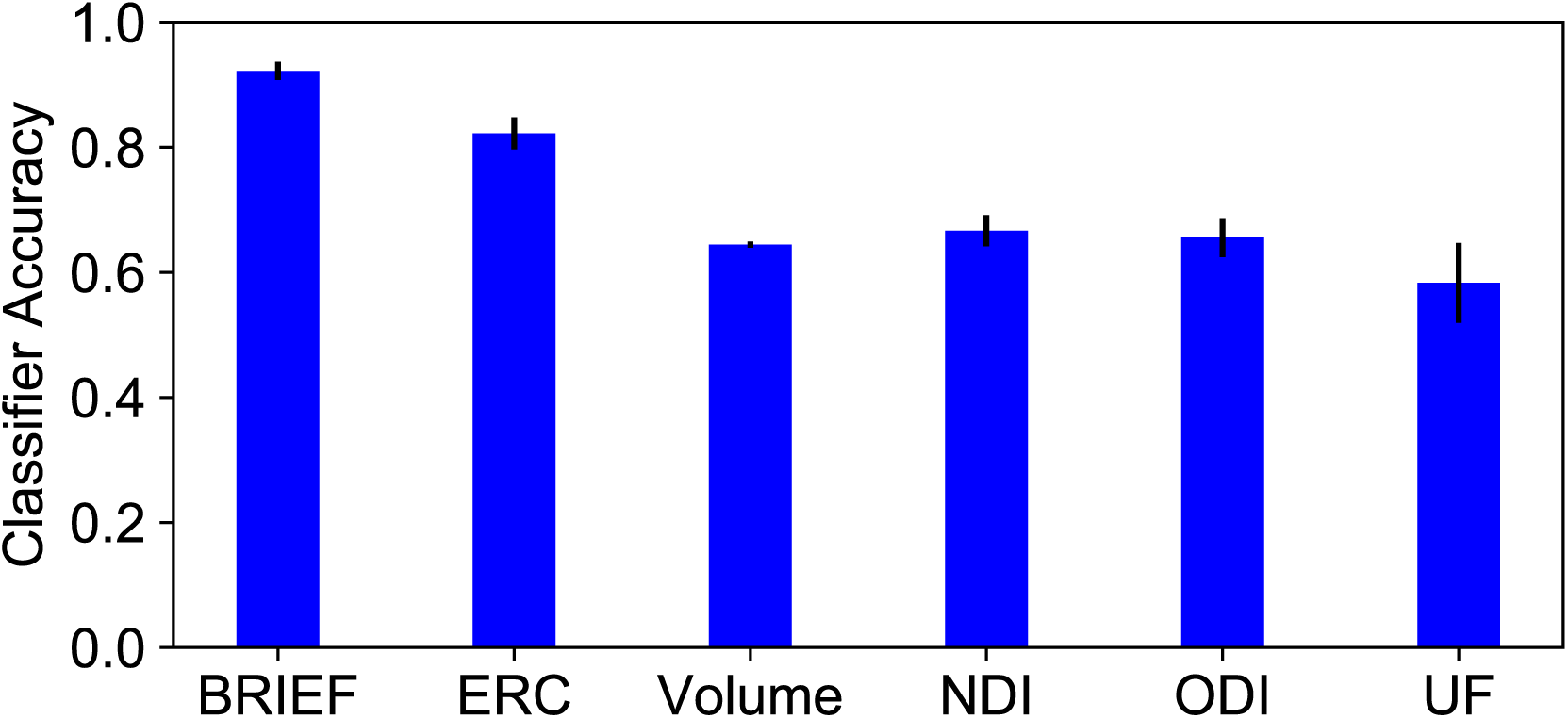
Classifier performance for predicting diagnostic category. Error bars indicate standard error of the mean classifier performance achieved across the five iterations employed.

**Figure 3.**
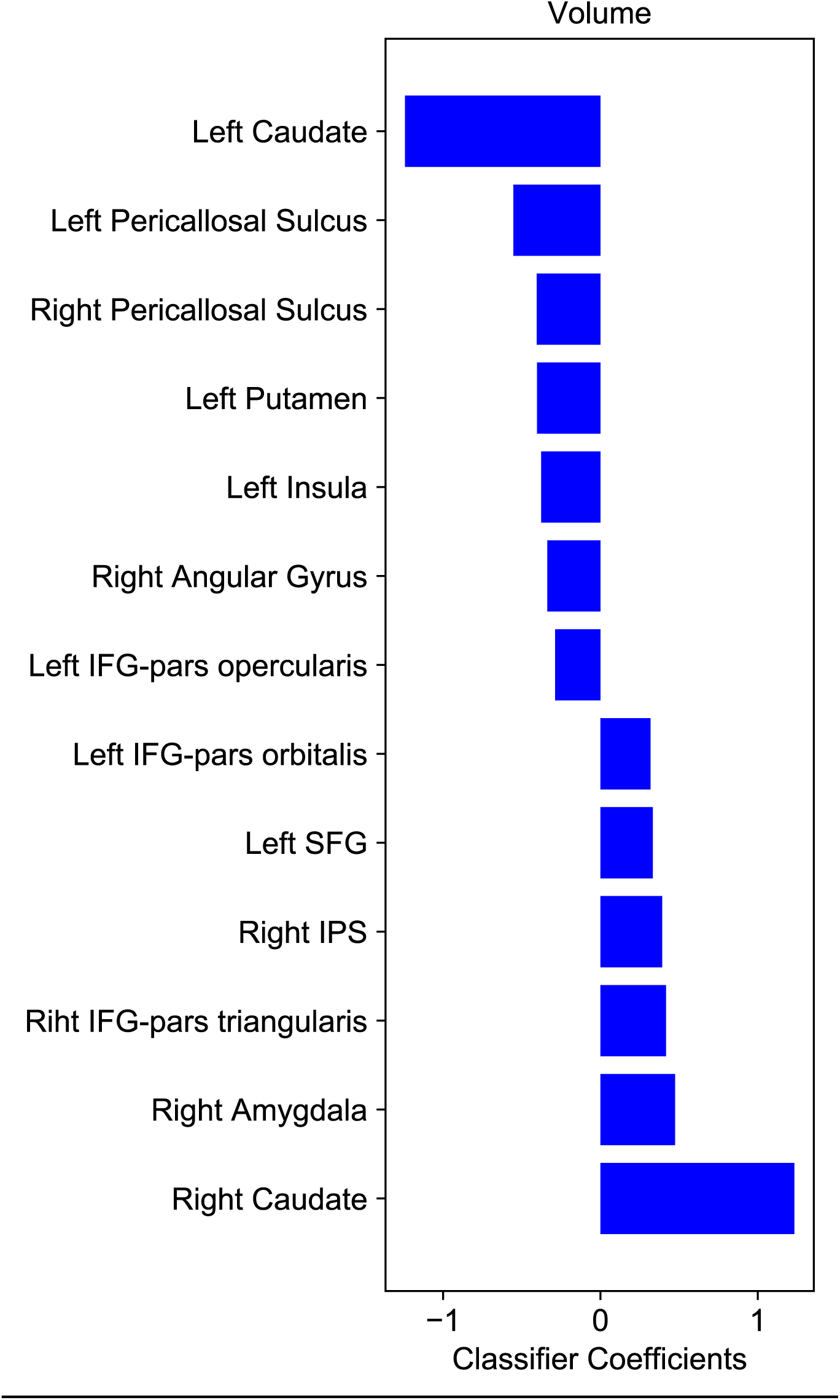
Selected regions of EF and ER for cortical and subcortical volume. Classifier coefficients indicate feature importance rankings of the selected regions for predicting diagnostic category.

**Figure 4.**
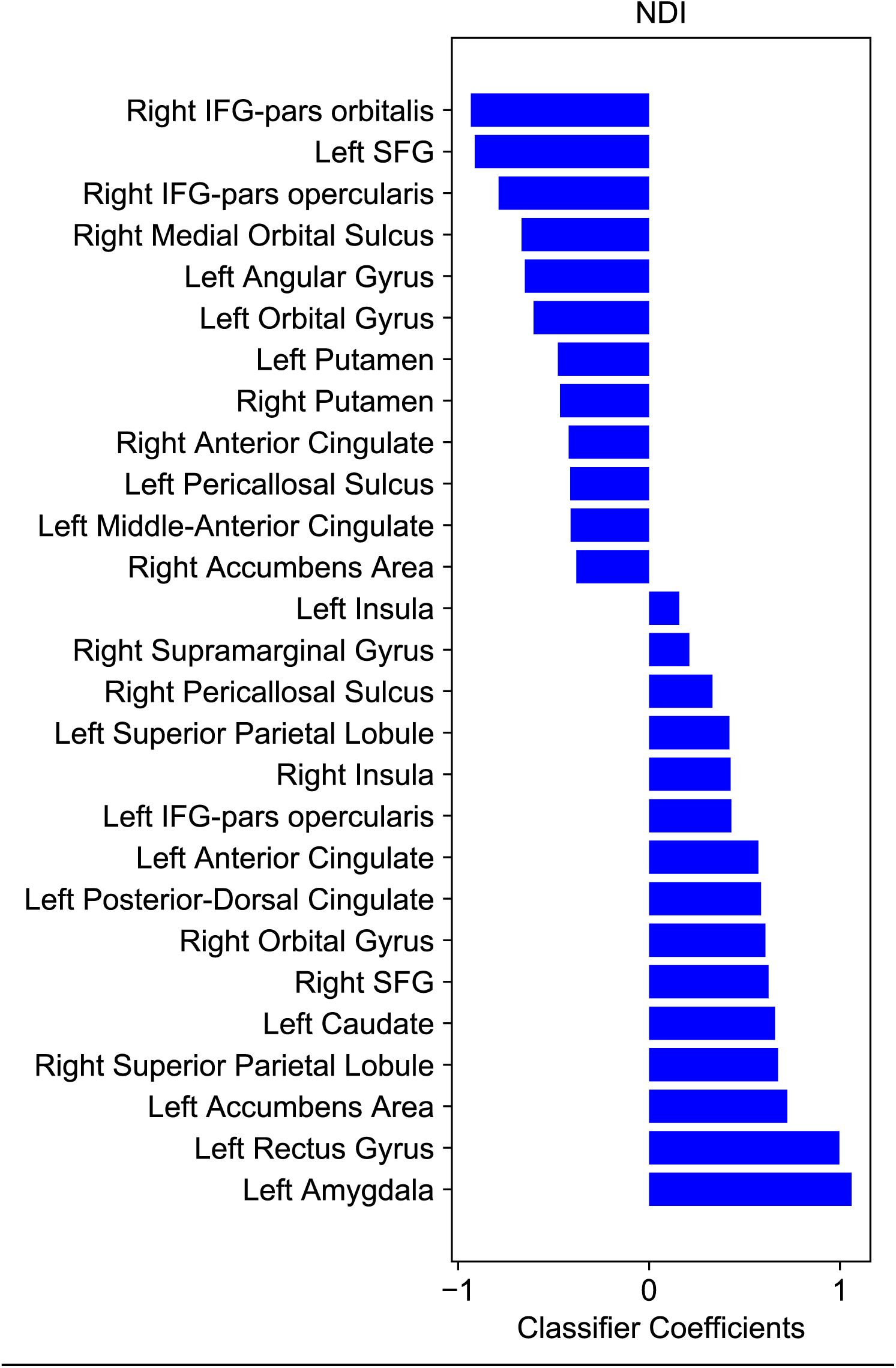
Selected regions of EF and ER for NDI metrics. Classifier coefficients indicate feature importance rankings of the selected regions for predicting diagnostic category.

**Figure 5.**
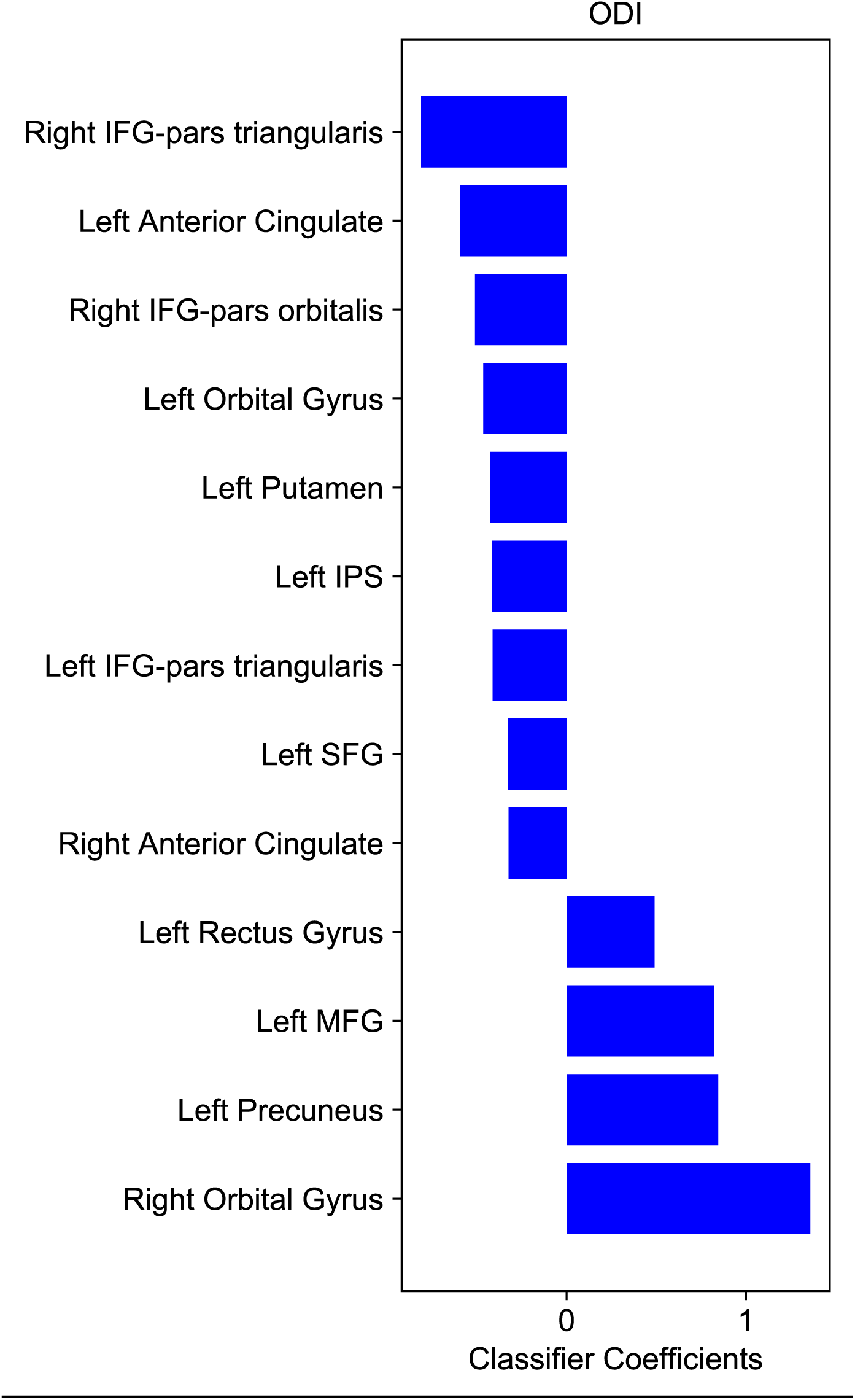
Selected regions of EF and ER for ODI metrics. Classifier coefficients indicate feature importance rankings of the selected regions for predicting diagnostic category.

The model including the teacher ratings of BRIEF MCI yielded the highest classifier performance, with an average accuracy of .922 (p < .001) across the five cross-validation indices. Teacher ratings of emotion regulation derived from the ERC reached an accuracy of .822 (p < .001). For our structural brain imaging measures of cortical and subcortical volume in EF and ER regions, the classifier achieved an average performance of .644 (p < .002). The classifier reached the following accuracy for the diffusion weighted imaging measures: .667 (p < .001) for NDI in EF and ER regions, .656 for ODI in EF and ER regions, .583 (p < .028) for fractional anisotropy in UF, .528 (p > .660) for the fractional anisotropy in FAT, and .522 (p > .610) for fractional anisotropy in cingulum. Thus, our machine learning approach has identified teacher ratings of EF and ER, as well as the corresponding neurobiological measures of cortical and subcortical volume, neurite density (both NDI and ODI), and fractional anisotropy in the UF pathway as significant predictors of ADHD diagnostic category.

### Predicting symptoms of hyperactive/impulsive behavior and inattention

Next, we sought to identify the degree to which our target measures contribute to dimensional constructs of ADHD, namely symptoms of hyperactive/impulsive behavior and inattention, assessing each set of the target measures (teacher ratings, structural imaging measures of volume, diffusion weighted imaging measures of NDI, ODI, and fractional anisotropy in the target fiber pathways). We ran separate linear regressions predicting hyperactivity and inattention symptomology, with these target measures entered into the model as predictors, with age and gender also included as covariates. For analyses that included neural measures as target predictors, we also included total intracranial volume and scan quality as covariates. For structural imaging measures of volume, this included the scan quality rating for the T1 scan. For diffusion imaging measures (NDI, ODI, and fractional anisotropy in fiber pathways), this included the number of directions kept, after automated quality control, for the diffusion-weighted scan. Below, we outline the measures that showed statistically reliable relationship with ADHD symptomology.

#### Teacher Ratings

Our analysis of the three teacher rating measures (BRIEF MCI, emotion regulation z score in ERC, and negativity *z*-score in ERC) indicated a reliable relationship between each target measure and symptoms of inattention and hyperactive/impulsive behavior. Specifically, BRIEF MCI ratings had a positive relationship with hyperactivite/impulsive behavior (*t*(174) = 4.*567*, p < .001) and inattention symptomology (*t*(174) = 5.583, p < .001). Similarly, the negativity *z*-score in the ERC also yielded a reliable positive relationship for both hyperactive/impulsive behavior (*t*(174) = 7.912, p < .001) and inattention (*t*(174) = 6.395, p < .048). The emotion regulation *z*-score measure from the ERC exhibited a negative relationship with symptoms of hyperactive/impulsive behavior (*t*(174) = -8.316, p < .001) and inattention (*t*(174) = -6.727, p < .001).

#### Structural Imaging Measures of Volume

Among the EF and ER regions identified by our RFE approach, the regions that were reliable predictors of both hyperactive/impulsive behavior and inattention symptoms were the left caudate (*t*(162) = -1.986, p < .049 for hyperactivity; *t*(162) = -2.677 , p < .008 for inattention), the left pericallosal sulcus (*t*(162) = -2.592, p < .010 for hyperactivity; *t*(162) = -2.014, p < .046 for inattention), the right inferior frontal gyrus – pars triangularis ( *t*(162) = 3.287, p < .001 for hyperactivity; *t*(162) = 2.794, p < .006 for inattention), and the right angular gyrus (*t*(162) = -3.642, p < .044 for hyperactivity; *t*(162) = -2.210, p < .029 for inattention). In addition to these regions that showed reliable relationship with both symptoms, the left inferior frontal gyrus – pars opercularis (*t*(162) = -2.198, p < .029) and the left inferior frontal gyrus – pars orbitalis were associated with symptoms of hyperactive/impulsive behavior (*t*(162) = 2.095, p < .038), while the left anterior segment of the circular sulcus of the insula (*t*(162) = -2.114, p < .036) and the right caudate (*t*(162) = 2.136, p < .034) were associated with symptoms of inattention.

#### Diffusion Weighted Imaging Measures of NDI and ODI

Among the EF and ER regions identified by our RFE approach, several regions further showed reliable relationship with both hyperactive/impulsive behavior and inattention symptoms of ADHD. For NDI, these regions were the left rectus gyrus (*t*(148) = 3.313, p < .001 for hyperactive/impulsive behavior; *t*(148) = 3.981, p < .001 for inattention) and the right inferior frontal gyrus – pars orbitalis (*t*(148) = -3.026, p < .003 for hyperactive/impulsive behavior; *t*(148) = -3.246, p < .001 for inattention). For ODI, these regions also included the left rectus gyrus (*t*(162) = 2.379, p < .019 for hyperactive/impulsive behavior; *t*(162) = 3.199, p < .002 for inattention) and the right inferior frontal gyrus – pars orbitalis (*t*(162) = -2.600, p < .010 for hyperactivity; *t*(162) = -3.644, p < .001 for inattention). ODI metrics further implicated the left anterior cingulate to predict both symptoms (*t*(162) = - 2.972, p < .003 for hyperactive/impulsive behavior; *t*(162) = -3.323, p < .001 for inattention). Additionally, NDI in the right anterior cingulate cortex exhibited a reliable relationship with symptoms of hyperactive/impulsive behavior (*t*(148) = -2.137, p < .034). Further regions that exhibited significant relationships with symptoms of inattention were the right superior frontal gyrus for NDI (*t*(148) = 2.080, p < .039), as well as the left middle frontal gyrus (*t*(162) = 2.018, p < .045) and bilateral inferior frontal gyri – pars triangularis (*t*(162) = -2.155, p < .033 for the left hemisphere, and *t*(162) = - 2.029, p < .044 for the right hemisphere) for ODI.

#### Diffusion Weighted Imaging Measures of FA in Fiber Pathways

Among the three pathways assessed (FAT, UF and the cingulum), our machine learning results had implicated the UF to be the only pathway that can significantly predict diagnostic category. Additional regression analyses assessing the relationship between fiber pathway FA and ADHD symptomology indicated no reliable/measurable relationship for any of the pathways evaluated.

#### Incremental benefit of the identified target measures in predicting ADHD symptomology

Upon identifying the target measures that can reliably predict both ADHD diagnostic category and symptomology, we next evaluated the incremental value across the teacher ratings and the implicated neurobiology in predicting ADHD symptomology. That is, does the implicated neurobiology of EF and ER have a benefit above and beyond teacher ratings of EF and ER in predicting ADHD symptomology? And if so, what are the regions that provide this additional benefit for each critical measure of ADHD symptomology? To this end, we assessed a full model that included all the significant measures identified above, and we further evaluated changes in *adjusted-R^2^* in our models as we add each critical set of predictors (see Table 1). We constructed two full models that included all the target measures that were identified to significantly predict both ADHD diagnostic category and symptom severity dimensionally for each of the symptoms of hyperactive/impulsive behavior and inattention associated with ADHD. For each analysis, we also included our control variables of age, gender, total intracranial volume, T1 scan quality, and DWI scan quality. Our goal was to evaluate which measures would still hold a statistically reliable relationship with ADHD symptomology when all the implicated measures were assessed conjointly in these full models.

**Table 1.** Incremental benefit among the implicated target measures in predicting ADHD symptoms of inattention and hyperactive/impulsive behavior. The table presents changes in adjusted-*R^2^* with the addition of 1) teacher ratings of EF and ER, and 2) the neurobiology of EF and ER that significantly predicted symptoms in our full model.

| **Model**<br>***Adjusted-R<sup>2</sup>*** | Demographics<br>(Age + child sex) | Demographics<br>and Ratings | Demographics, Ratings<br>and Neurobiology |
| --- | --- | --- | --- |
| Inattention | .004 | .433 | .454 |
| Hyperactive/impulsive | .005 | .452 | .507 |

For symptoms of hyperactive/impulsive behavior, all teacher variables maintained a reliable relationship (*t*(159) = 3.478, p < .001 for BRIEF EMC t scores, *t*(159) = 5.971, p < .001 for ERC z scores of negativity, and *t*(159) = -6.279, p < .001 for ERC z scores of emotion regulation). Additionally, neurobiological measures that were still reliable were the left pericallosal sulcus volume (*t*(159) = -2.567, p < .011), the left inferior frontal gyrus – pars orbitalis volume (*t*(159) = 3.026, p < .003), NDI in the right inferior frontal gyrus – pars orbitalis (*t*(159) = -2.159, p < .032), and ODI in the left anterior cingulate cortex (*t*(159) = -2.222, p < .028).

For symptoms of inattention, the target variables that remained statistically reliable were BRIEF EMC (*t*(155) = 4.952, p < .001), ERC negativity (*t*(155) = 3.833, p < .001), ERC emotion regulation (*t*(155) = -4.060, p < .001), left caudate volume (*t*(155) = -2.166, p < .032), and NDI in the right inferior frontal gyrus – pars orbitalis (*t*(155) = - 2.419 , p < .017).

Thus, when considering the additional benefit of neurobiological measures for predicting ADHD symptomology above and beyond the target teacher rating measures of EF and ER, our results implicate the right inferior frontal gyrus (specifically the pars orbitalis) as a region that predicts for both symptoms of inattention and hyperactive/impulsive behavior, the left caudate for symptoms of inattention, as well as the left pericallosal sulcus, the left inferior frontal gyrus and the left anterior cingulate cortex as additional regions associated with hyperactive/impulsive behavior.

### Addressing comorbid ODD in the ADHD sample

So far we have assessed our main goal of identifying the relative importance of our target measures for predicting ADHD diagnostic category and ADHD symptomology of hyperactivite/impulsive and inattention. Recall that our ADHD sample has 69.4% comorbid ODD. Thus, when considering the overall clinical utility of the target measures assessed in the current study, an important question arises regarding if they have any further predictive utility for differentiating comorbid ODD in the ADHD sample. To address this question, we ran additional models that assess the degree to which a classifier can successfully predict the presence of comorbid ODD in the ADHD sample. Accordingly, using the same modeling protocol above, a classifier was trained to distinguish between ADHD only versus ADHD+ODD diagnosis in our ADHD sample, using the same target measures employed for the analyses reported above.

#### Teacher Ratings

Two models were trained to assess the performance of the classifier in predicting comorbid ODD diagnosis based on the BRIEF teacher ratings of executive function, and ERC teacher ratings of emotion regulation and negativity. Among the two models assessed, only the ERC model achieved a reliable classifier performance, with an average accuracy of .741 (p < .001). Accordingly, our findings indicate that while a strong predictor of ADHD diagnostic category and symptomology, BRIEF ratings of executive function might not have predictive utility in further distinguishing comorbid ODD diagnosis within the ADHD sample. ERC ratings on the other hand might be a more suitable indicator of comorbid ODD diagnosis.

#### Neural Measures

We ran six models to assess the predictive utility of our neural measures in predicting comorbid ODD diagnosis. These models each evaluated the selected regions for structural imaging measures of volume, DWI measures of NDI, and DWI measures of ODI, identified for predicting ADHD diagnostic category reported above, as well as the fractional anisotropy in the target fiber pathways, namely FAT, cingulum, and UF. None of these models were able to reliably predict comorbid ODD within our ADHD sample. Thus, our findings indicate that the implicated neurobiology important for predicting ADHD diagnostic category did not have further predictive utility in predicting comorbid ODD diagnosis.

## DISCUSSION

The current investigation provided a unique assessment of the neurobiology of ADHD in comparison with teacher ratings of EF and ER in a very young age range (ages 4-to-7), as they relate to predicting ADHD diagnostic category and ADHD symptomology of hyperactive/impulsive behavior and inattention. We used machine learning to evaluate structural brain imaging measures, as well as diffusion weighted imaging measures of neurite density and fiber pathway fractional anisotropy as they relate to ADHD. Teacher ratings of EF were the most robust predictor of ADHD diagnostic category while teacher ratings of ER were more critical for predicting comorbid ODD. When considering the additional benefit of neurobiological measures for predicting ADHD symptomology above and beyond the target teacher rating measures of EF and ER, our results implicate the right inferior frontal gyrus (specifically the pars orbitalis) as a region that predicts for both symptoms of inattention and hyperactive/impulsive behavior, the left caudate for symptoms of inattention, as well as the left pericallosal sulcus, the left inferior frontal gyrus and the left anterior cingulate cortex as additional regions associated with hyperactive/impulsive behavior. Interestly, our neural measures implicated for predicting ADHD diagnostic category had no further incremental validity in predicting comorbid ODD symptoms.

Below, we further summarize our findings and discuss their clinical and theoretical implications.

### Teacher ratings of executive function and emotion regulation

In the current study we further examined the neurobiology of ADHD, and also determined the extent to which such neural measures may be useful in predicting ADHD symptomology and comorbid ODD, above and beyond teacher ratings of EF and ER. Consistent with recent work from our group (Oztekin et al., 2021), teacher ratings of EF were the most dominant predictor of ADHD diagnostic category, but not comorbid ODD. Teacher ratings from the ERC were also a reliable predictor of ADHD diagnostic category, although not as high as teacher ratings of EF. In contrast to BRIEF ratings, ratings of ERC could reliably predict comorbid ODD. Thus, while our investigation points to teacher ratings of EF as the most diagnostic predictor of ADHD diagnostic category, teacher ratings of ER stand out as a more critical predictor for comorbid ODD diagnosis in this very young age range. Consistent with emerging work suggesting ER as critical component of ADHD (Graziano & Garcia, 2016; Shaw et al., 2014), our results suggest that early intervention efforts should target children’s ER skills (rather than EF) given its relation to both core ADHD symptomlogy and ODD. Targetting ER skills during the preschool period is especially important given the high comorbidity of ADHD and ODD during this period (Harvey et al., 2016).

### Neurobiology of ADHD, its symptomology and their incremental benefit beyond teacher ratings of EF and ER

With respect to evaluating the neurobiology of EF and ER for ADHD, our analytical approach followed three steps. Initially, machine learning models were run to predict ADHD diagnostic category in our sample. Then, the target measures were further evaluated to assess whether they show reliable associations with dimensional assessments of ADHD symptomology, namely symptoms of inattention and hyperactive/impulsive behavior. Third, among the target measures that exhibited reliable associations with ADHD in these two steps, we further assessed their relative utility in comparison to the teacher ratings of EF and ER. Our overarching goal was to identify the critical neurobiology that provided diagnostic utility above and beyond those provided by the target teacher ratings alone. We next discuss the key neurobiology that were identified in this three-step approach.

The inferior frontal gyrus was a prominant brain region identified in our current investigation. Specifically, microstructure in this region was associated with both symptoms of ADHD. Prior cognitive neuroscience research has consistently implicated this region as critical for successful and efficient resolution of interference in working memory. Numerous neuroimaging studies have noted enhanced neural activation in this region in the presence of interference (Badre & Wagner, 2005; D’Esposito et al., 1999; Jonides & Nee, 2006; Oztekin & Badre, 2011; Oztekin et al., 2009). The critical role of this region in supporting the control mechanisms that resolve interference in working memory has been further established by studies demonstrating that patients with lesion in this area are more susceptible to making errors in the face of interference (Feredoes et al., 2006; Thompson-Schill et al., 2002), and by repetitive transcranial magnetic stimulation evidence indicating that inhibition of this region increases error rates in the presence of interference (Feredoes et al., 2006).

Another relevant and related context that implicates the right inferior frontal cortex has been inhibitory control operations identified in Go-No Go paradigms such as the Stop Signal Task (Hannah & Aron, 2021). Notably, previous research has identified this region as a potential modulatory neural measure for ADHD-related performance deficits in this task (Chevrier & Schachar, 2020; Tremblay et al., 2020). Given that inhibitory control and working memory deficits are commonly observed in ADHD (Hammer et al., 2015; Karalunas et al., 2017; Palladino & Ferrari, 2013; Raiker et al., 2019; Raiker et al., 2012), it will be important for future research to clarify whether its potential role in ADHD is via its modulatory role in controlled working memory processes.

In addition to the inferior frontal gyrus, our study further implicated the left caudate for symptoms of inattention, as well as the left anterior cingulate cortex, and the left pericallosal sulcus as critical neural measures for symptoms of hyperactive/impulsive behavior. The further implication of these regions of the emotion regulation network is consistent with the contention that EF related effects might not predominantly emerge in this very young age rage (Halperin & Schulz, 2006), and extends prior research that have identified important contributions from subcortical volume measures (Hoogman et al., 2020; Rosch et al., 2018). Thus, with respect to behavioral/cognitive and neural measures of theoretical importance for ADHD, our findings further stress the importance of conjointly evaluating EF and ER.

### Critical Future Directions

With respect to the neurobiology of ADHD, the contributions of individual differences in white matter and neuronal microstructure gleaned from diffusion weighted imaging have been notably understudied. Studies exploring the feasibility of machine learning for classification of ADHD have mostly focused on T1-weighted structural data, for instance focusing on cortical thickness measures (Colby et al., 2012; Ghiassian et al., 2016; Oztekin et al., 2021; Peng et al., 2013; Qureshi et al., 2017; Sen et al., 2018), with the majority of studies focusing on functional MRI, resting-state data (Colby et al., 2012; Dai et al., 2012; Du et al., 2016; Ghiassian et al., 2016; Qureshi et al., 2016; Qureshi et al., 2017; Sen et al., 2018; Sidhu et al., 2012; Wang et al., 2017), functional brain volumes (Tan et al., 2017) or task-based functional data (Hart et al., 2014). Accordingly, a major criticism has been the issue of transportability (Foster et al., 2014; Woo et al., 2017), and that the findings do not have the potential to generalize to or be easily applied in clinical settings. Because DWI metrics can be obtained in a short anatomical scan, they have better promise for clinical settings. In addition to their potential clinical utility, our findings critically implicate their theoretical importance for ADHD. Most importantly, our current findings implicate neurite density as an important neural measure that could jointly predict ADHD diagnostic category and its symptomology. As such, the current set of findings strongly implicate the critical importance of pursuing diffusion weighted imaging measures with respect to the underlying neurobiology and potential functional impairments associated with ADHD.

It is also important to note how the neural measures identified for predicting ADHD diagnostic category and the associated symptoms could not further predict comorbid ODD. This result is consistent with previous meta-analyses analyses that found no association between brain dysfunctions and comorbid conditions in ADHD (Cortese et al., 2012; Hart et al., 2012), and previous studies that have observed differential neurobiology for ADHD and CC/ODD (reviewed in Rubia, 2011). These findings are in line with prior work suggesting that ADHD has a stronger neurobiological basis whereas ODD may have stronger ties to environmental/family risk factors (Rowe et al., 2002). These results further support early intervention guidelines by the American Academy of Pediatrics (Wolraich et al., 2019) that recommend behavioral parent training as a first line of treatment for preschoolers with ADHD, especially given the high comorbidity rates with ODD. Nevertheless, it will be important to further evaluate these neural markers to determine their utility in predicting not only the developmental course of ADHD but also children’s response to both medical and psychosocial interventions.

## Data Availability

Data available on request due to privacy/ethical restrictions

## Conflict of Interest

None reported

## Acknowledgements

The AHEAD study is funded by grants from the National Institute of Health (R56MH108616, R01MH112588 and R01DK119814) to P. A. Graziano and A. S. Dick. This research was supported by a Computational Administrative Supplement to parent grant R01MH112588.

## Data Availability

Data available on request due to privacy/ethical restrictions.

---

**Supplementary Table 1.** Performance metrics for predicting ADHD diagnostic category for whole brain measures.

| **Models** | Volume | Area | CT | Curvature | NDI | ODI |
| --- | --- | --- | --- | --- | --- | --- |
| Accuracy | .844 | .683 | .683 | .683 | .739 | .794 |
| Permutation<br>p value | .001 | .001 | .001 | .001 | .001 | .001 |
| F<sub>1</sub> | .837 | .667 | .654 | .633 | .720 | .773 |

Note. CT = Cortical thickness; NDI = Neurite Density Index; ODI = Orientation Dispersion Index

## Notes

### Competing Interest Statement

The authors have declared no competing interest.

### Author Declarations

The study was approved by The Health Sciences Institutional Review Board at the Office of Research Integrity at Florida International University, located in Miami, FL.

